# Potential Association of Mitochondrial Haplogroups and A8860G Mutation with Breast Cancer Risk

**DOI:** 10.1101/2021.02.12.21249541

**Authors:** Han N. Mohammed Fadhl, Farhad M. Abdulkarim

**Author notes:** Corresponding author (FMA). Contributionship: (HNM) and (FMA).

## Abstract

The last decade has witnessed great progresses regarding the molecular basis of breast cancer with discovery of different nuclear susceptibility genes; in addition investigations and researches regarding mitochondrial DNA (mtDNA) mutations in breast cancer have been started. Mitochondrial haplogroup determinants (single nucleotide polymorphism SNP) and somatic mitochondrial mutations have recently been studied as possible risk factors for carcinogenic processes in different tissues, hence in order to identify breast cancer related SNPs and haplogroups among the population of Sulaimaniyah city/Iraq, the entire mitochondrial genome of 20-breast cancer samples and comparable controls were sequenced. Haplogrep 2.0 was used for haplogroup identification; Chi-square and Fishers exact test were applied to assess relational significance. HV haplogroup in the cancer samples appeared to be a risk factor for breast cancer compared to the most common H haplogroup in control samples with a p-values of 0.002 and 0.006 respectively and an Odd Ratio (OR) = 28.00. Besides, SNP (A8860G) was also identified as a risk factor for breast cancer as compared to other randomly selected SNPs (A750G, A1438G and C7028T) with p values □0.05 and OR >1.

## 1. Introduction

Breast cancer is the most frequently diagnosed cancer in women [1]; representing a heterogeneous, debilitating disease of a multistep carcinogenic background [2]. The incidence of breast cancer in Kurdish women in north of Iraq is the same as in western countries and even higher especially in the young age group [3, 4]. Although, major progresses have been made over the past decade in exploring the molecular basis of breast cancer and several susceptibility nuclear-genes of high, moderate and low penetrance have been identified as predisposing factors for breast cancer, still these genes can explain only 10-15% of breast cancer cases [5, 6, and 7]. Recently, many attempts have been made to study possible relation of mitochondrial genome mutations with incidence of breast cancer and other types of cancer, including mitochondrial haplogroups that are determinants of a number of single nucleotide polymorphisms (SNP)’s gained by geographical and environmental impacts during the process of evolution throughout history [8]. To date several studies in different populations have been performed, figuring out significant relation between specific haplogroups and incidence of different cancers, including breast cancer [9, 10, 11, and 12] as well as identification of specific mutational variants in association with cancer cases in general as T16189C, G10398A and the deletion of mtDNA 4977 [13]. Mitochondrial DNA (mtDNA) is a circular molecule of 16569 bp, coding for two 2 ribosomal RNAs (12S and 16S), 22 transfer RNAs and 13 essential protein subunits of the oxidative phosphorylation system (OXPHOS) [14]. It is well known that (mtDNA) has a mutation rate several times higher than nuclear DNA [15], due to its limited repair mechanisms, lack of protective histones and its close proximity to the electron transport chain, which continuously generates free radicals [16]; in addition, mtDNA is organized in an economic pattern with its genes lacking introns [17]. It was in 1956, when Otto Warburg observed that cancers ferment glucose in the presence of oxygen, proposing that abnormalities in mitochondrial respiration may be responsible for cancer production [16, 18, and 19]. Now it’s well-established that changes in the biochemical processes that accompany the process of carcinogenesis as the aerobic glycolesis do not impair mitochondrial function but are rather essential for cancer cell viability [15].

The current study was performed to identify possible relation of specific haplogroups with incidence of breast cancer in the population of Sulaimaniyah city (a Kurd ethnic occupant city in north of Iraq), as well as recognition of significantly common SNPs among the breast cancer cases in the studied population, considering no such previous studies have been performed neither in the city nor in the locality

## 2. Materials and methods

### Sample selection

The study was approved by the Ethical Committee of Collage of Medicine/University of Sulaimani (Number 44, on January 30, 2017) and written consents were obtained from all participants. A total of 40 subjects (20 breast cancer tissue and 20 control samples) were recruited for the study. Control samples were taken from benign breast tissue specimens (fibroadenoma and non-proliferative fibrocystic breast disease). Breast cancer tissue samples were taken from mastectomy specimens of subjects already diagnosed as having invasive ductal carcinoma (Grade II and III) by core biopsy and with no family history of breast cancer. All samples were from unrelated individuals of the Kurdish ethnic population from the center of Sulaymaniyah city.

### DNA extraction

Total genomic DNA was extracted using DNA extraction kit (GeNet bio/South Korea). The extraction was performed according to the manufacturer’s instruction. Purity and concentration of the extracted DNA were obtained using a Biophotometer (Eppendorf/Germany).

### PCR amplification and sequencing

The entire mitochondrial genome was amplified in the form of four overlapping PCR fragments using long Taq kit (Dongsheng Biotech/China) and the primers listed in (Table 1).

**Table 1:**
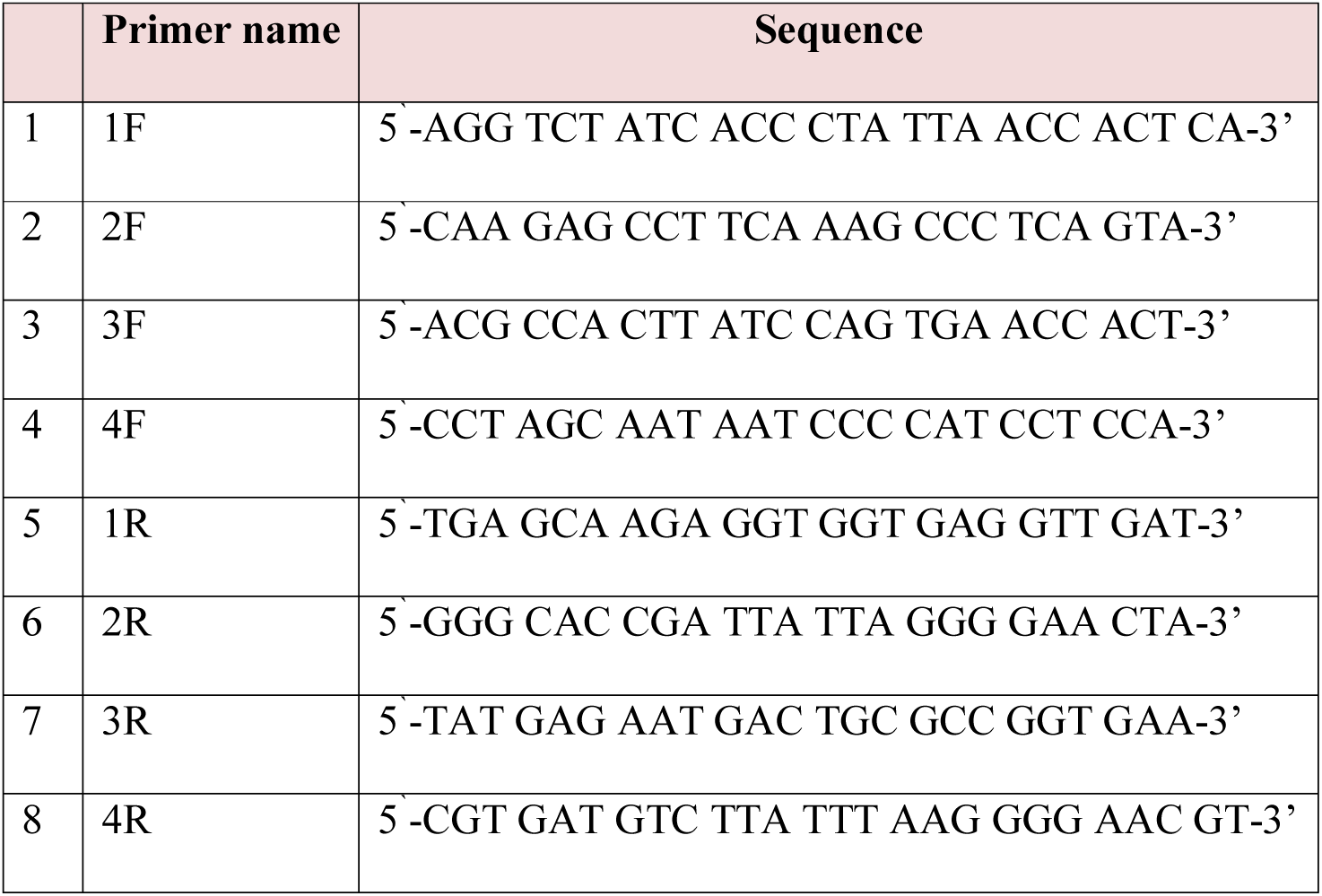
Sequence of the amplification Primers of four overlapping mitochondrial DNA fragments.

The amplified PCR products were purified using PCR purification kit (NORGEN biotek/Canada) and the primers listed in (Table 2) were used for sequencing of the amplified mtDNA fragments.

**Table 2:**
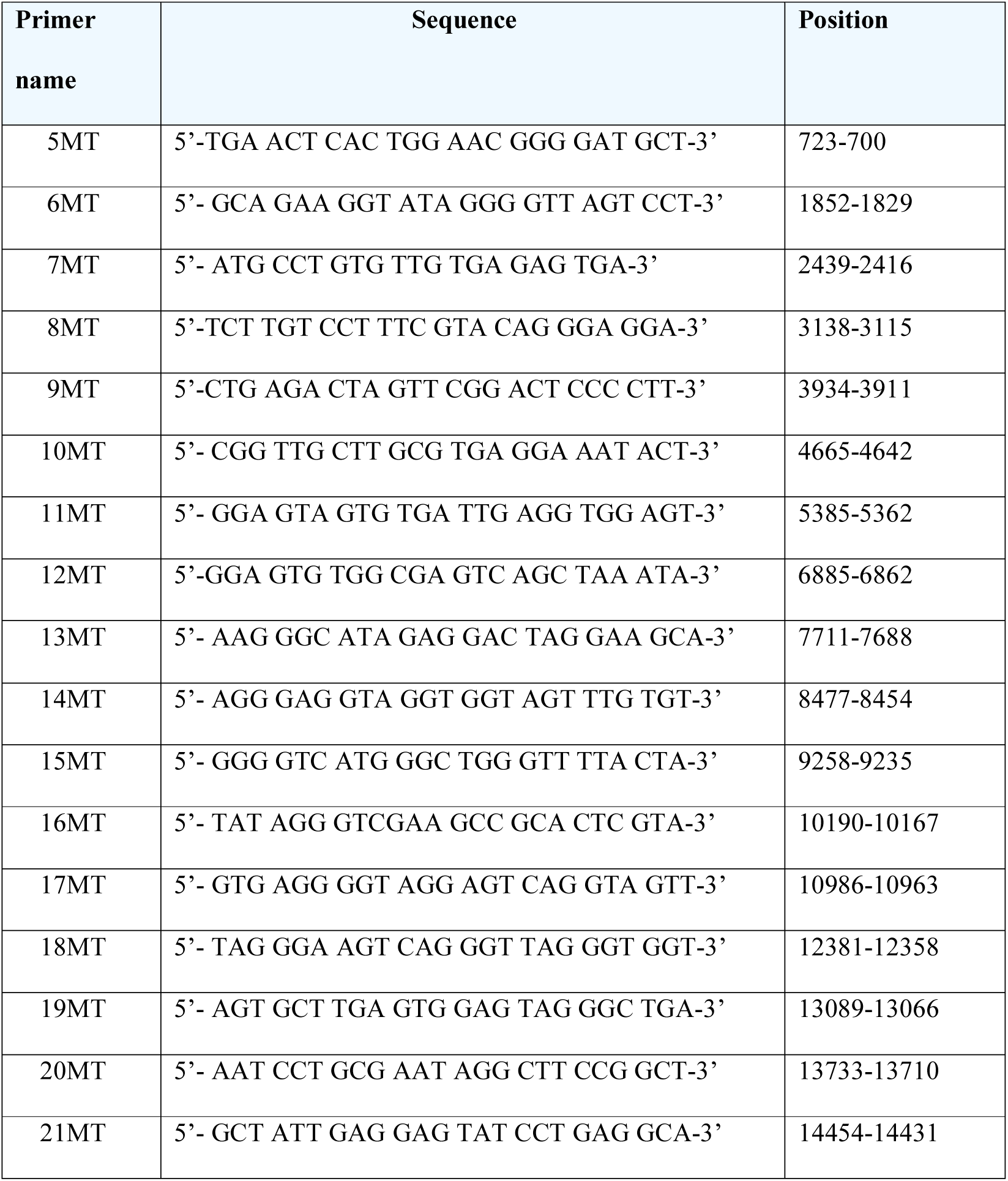
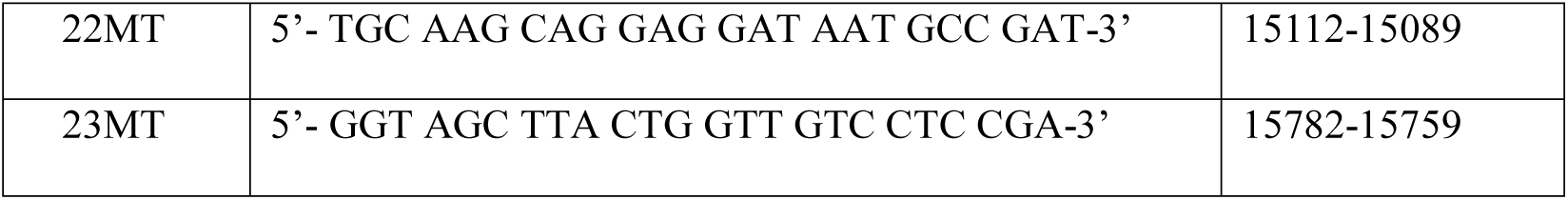
The Sequencing primers of the four overlapping fragments of mitochondrial DNA.

### Data analysis

The algorithm implemented in the HaploGrep 2.0 was used for identification of haplogroups [20]. Chi-square and Fishers exact test were used to determine the significance of relations of breast cancer with haplogroups and SNPs. The https://www.mitomap.org/MITOMAP website which provides a comprehensive database for human mitochondrial DNA was used for allocation of mutations, identifying types of the mutations and determining amino acid substitutions.

## 3. Results

A total of 344 mutations in the cancer samples and 203 mutations in the control samples were identified in the current study. The majority of the mutations were point mutations with only four insertion mutation regions. Based on the MITOMAP databases, single nucleotide polymorphisms (SNP) of the Kurdish ethnicity were identified and accounted for 74% of all mutations in the breast cancer samples, of which 61% were distributed in the coding region and 39% were in the non-coding region (Table 3). Whereas, in the control samples SNPs constituted 90% and the pattern of distributions were 58% in the coding region and 42% in the non-coding region (Table 4).

**Table 3:**
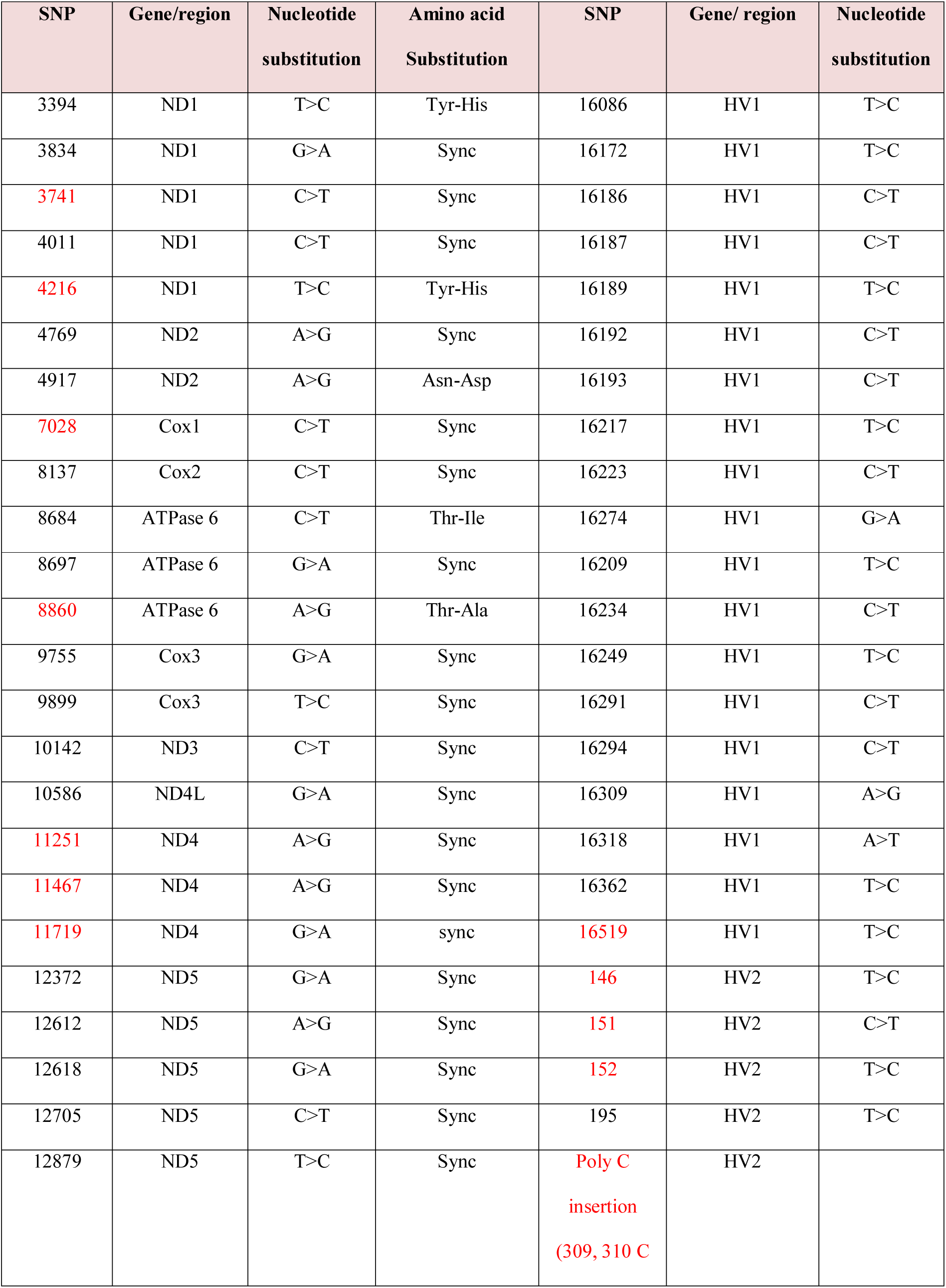
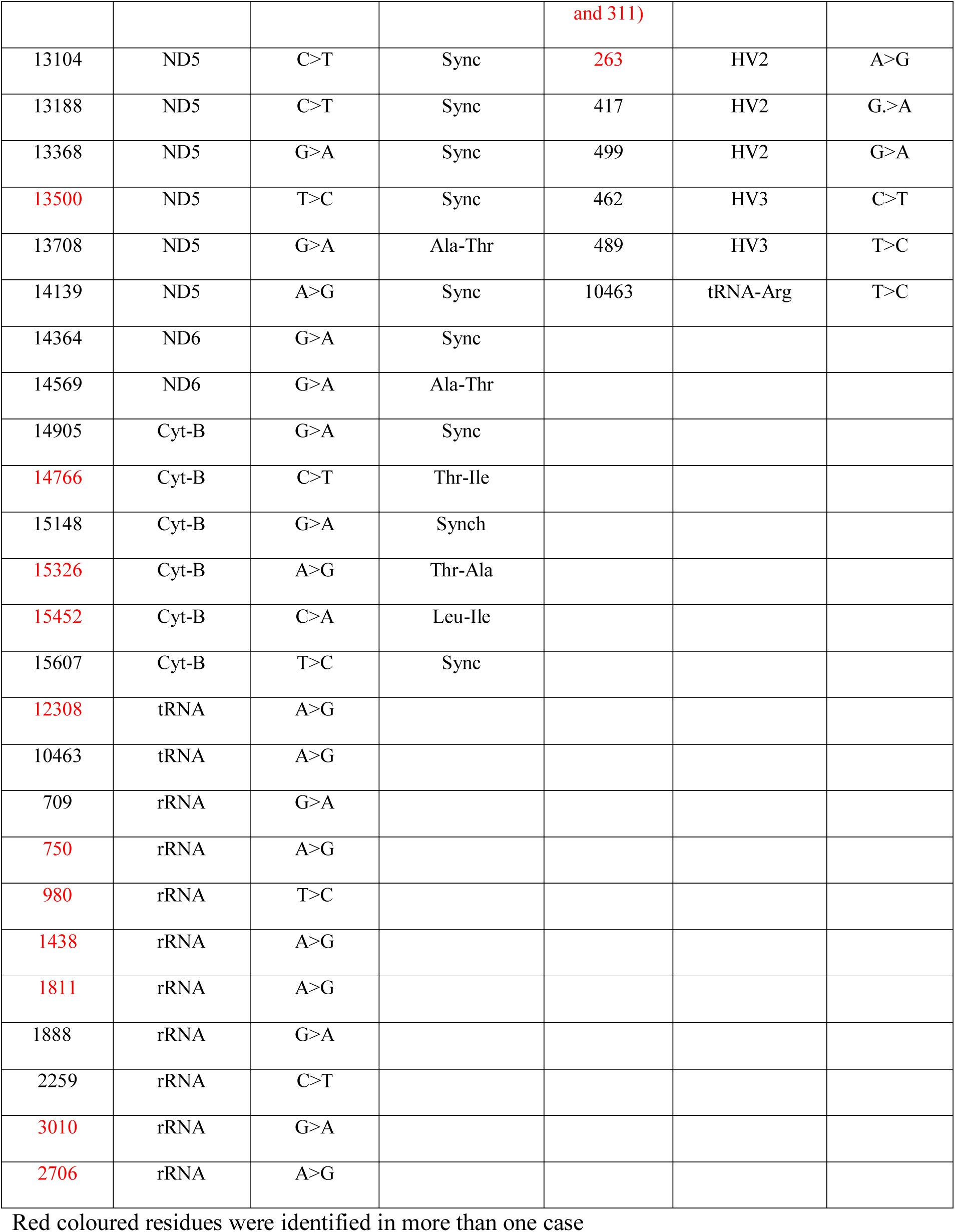
Total single nucleotide polymorphism (SNP) mutations in breast cancer samples.

**Table 4:** Total single nucleotide polymorphism (SNP) mutations in control group samples.

| SNP | Gene/region | Nucleotide substitution | a.a substitution | SNP | Gene/region | Nucleotide substitution | a.a substitution |
| --- | --- | --- | --- | --- | --- | --- | --- |
| 73 | HV2 | A>G | - | 1811 | rRNA | A>G |  |
| 146 | HV2 | T>C | - | 1438 | rRNA | A>G |  |
| 152 | HV2 | T>C | - | 2706 | rRNA | A>G |  |
| 189 | HV2 | A>G | - | 3010 | rRNA | G>A |  |
| 195 | HV2 | T>C | - | 4216 | ND1 | T>C | Tyr-His |
| 204 | HV2 | T>C | - | 4769 | ND2 | A>G | Synch, Met |
| 263 | HV2 | A>G | - | 5046 | ND2 | G>A | Val-Ile |
| 295 | HV2 | C>T | - | 5460 | ND2 | G>A | Ala-Thr |
| Poly C insertion (309, 310 C and 311) |  |  | - | 7028 | Cox1 | C>T | Synch |
|  |  |  | - | 7521 | tRNA | G>A |  |
| 462 | HV3 | C>T | - | 7789 | Cox2 | G>A | Synch |
| 489 | HV3 | T>C | - | 8860 | ATPase6 | A>G | Thr-Ala |
| 709 | rRNA | G>A | - | 8994 | ATP6 | G>A | Synch |
| 750 | rRNA | A>G | - | 11674 | ND4 | C>T | Synch |
| 1243 | rRNA | T<C |  | 11719 | ND4 | G>A | Sync |
| 13145 | ND5 | G>A | Ser-Asn | 12705 | ND5 | C>T | Synch |
| 14766 | Cyt-B | C>T | Thr-Ile |  |  |  |  |
| 15326 | Cyt-B | A>G | Thr-Arg |  |  |  |  |
| 16189 | HV1 | T>C |  |  |  |  |  |
| 16218 | HV1 | C>T |  |  |  |  |  |
| 16223 |  | C>T |  |  |  |  |  |
| 16519 | HV1 | T>C |  |  |  |  |  |

Red coloured residues were identified in more than one case

Nine Western Eurasian haplogroups and their subclasses were identified in both cancer and control samples using the Haplogrep 2.0 program. Haplogroups: HV, N, R, U, J, T and H were identified in breast cancer samples, on the other hand H, HV, N, R0, J, X and W haplogroups were identified in the control samples. The most common haplogroup in the control samples was the H-haplogroup (60%), while in breast cancer samples the frequency was less common (5%). In contrast, commonly occurring haplogroup in breast cancer samples was HV (35%) followed by N (25%) (Table 5 A & B).

**Table 5:** The identified Haplogroups in breast cancer (A) and control (B) subjects.

| Breast cancer samples |  |  | Control samples |  |  |
| --- | --- | --- | --- | --- | --- |
| Haplogroups | Frequency of occurrence | % | Haplogroups | Frequency of occurrence | % |
| HV | 7 | 35% | H | 12 | 60% |
| N | 5 | 25% | HV | 3 | 20% |
| U7 | 2 | 10% | N | 1 | 5% |
| R0 | 2 | 10% | R0 | 1 | 5% |
| J | 1 | 5% | J1 | 1 | 5% |
| U1 | 1 | 5% | X | 1 | 5% |
| T | 1 | 5% | W | 1 | 5% |
| H | 1 | 5% | - | - | - |
| Total | 20 | 100% | Total | 20 | 100% |

A statistically significant association between haplogroup HV and breast cancer was identified using Chi-square and Fisher’s exact tests, where the p-values were 0.002 and 0.006, respectively, and the odd ratio (OR) = 28, indicating that the HV haplogroup is a high risk factor for the incidence of breast cancer.

Furthermore, the homoplasmic mutation, SNP (A8860G) (Figure 1) was identified in all 20 breast cancer samples (100%), while in control samples it was less frequent and identified in only 4 samples (20%).

**Figure 1:**
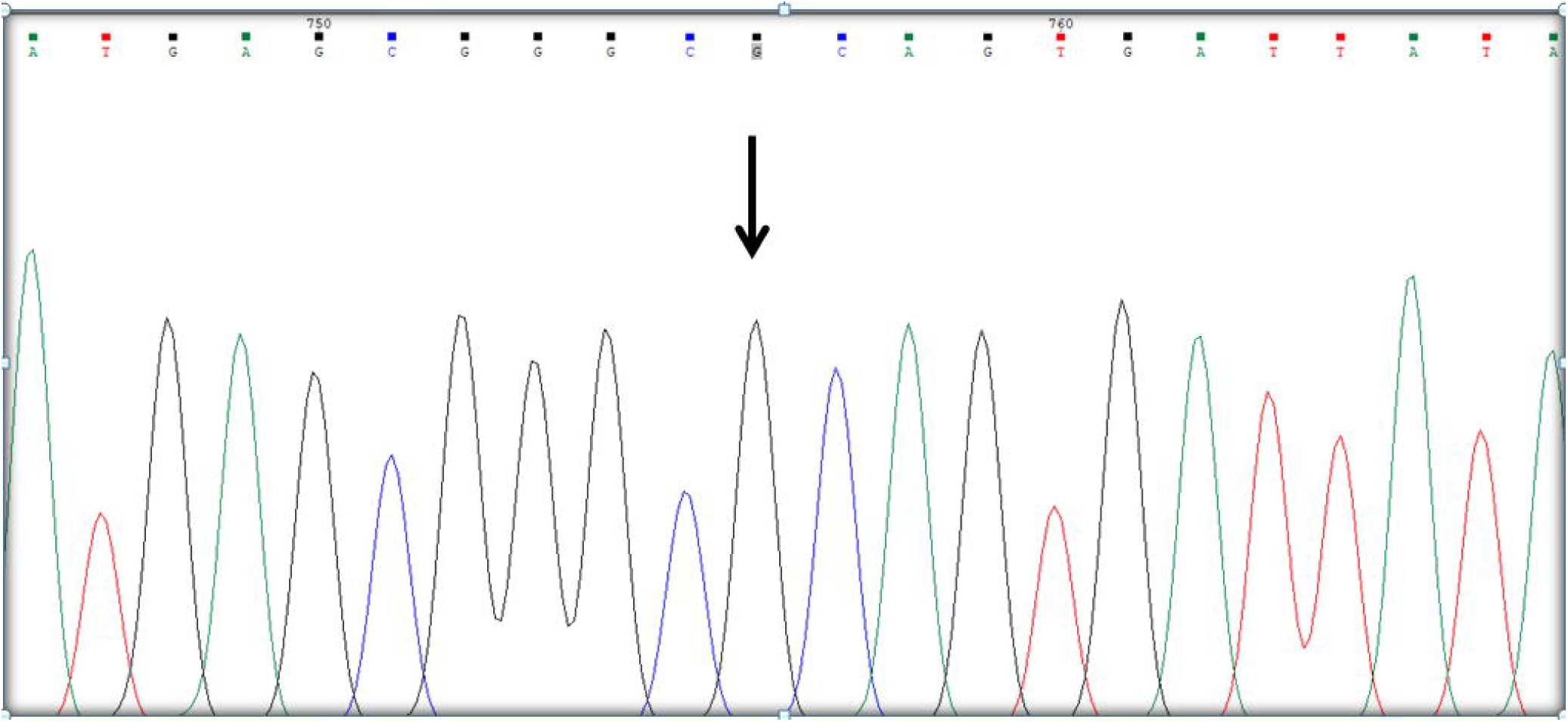
Electropherogram and sequence of the A8860G region. Point mutation site is indicated by an arrow

To identify the significance of this mutation, Chi-square, Fishers exact test were used and OR were calculated and compared with three other randomly selected SNPs (A750G, A1438G and C7028T) (Figure 2a, 2b and 2c).

**Figure 2a:**
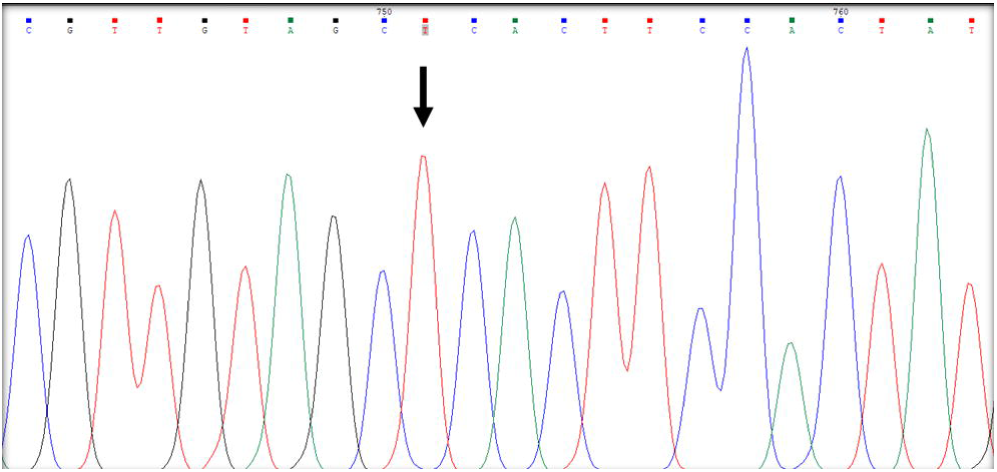
Electropherogram and sequence of the A750G region. Point mutation site is indicated by an arrow

**Figure 2b:**
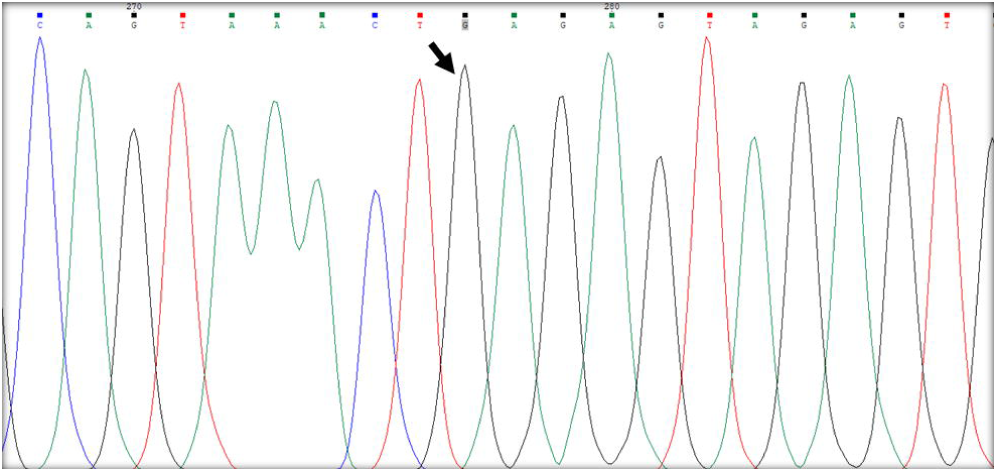
Electropherogram and sequence of the A1438G region. Point mutation site is indicated by an arrow

**Figure 2c:**
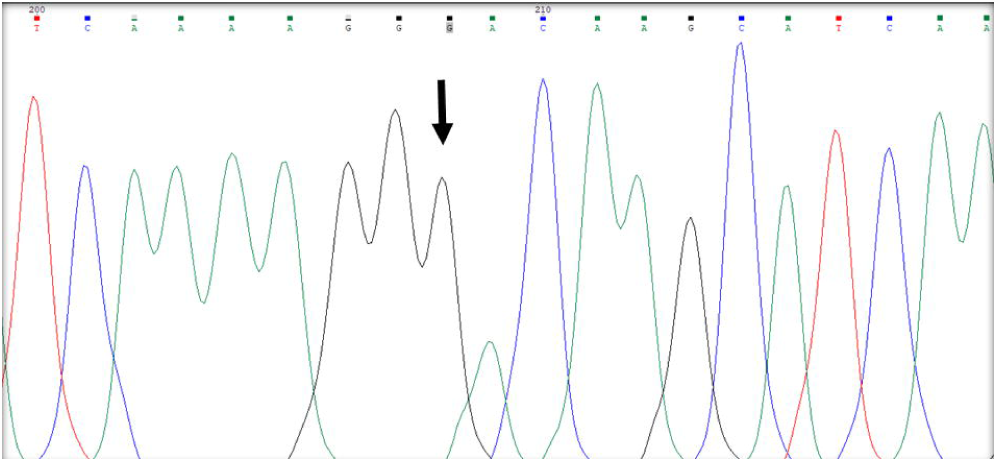
Electropherogram and sequence of the C7028T region. Point mutation site is indicated by an arrow

As indicated in (Table 6), the OR values were greater than 1 and the p-values were less than 1, indicating SNP (A8860G) as a risk factor for developing breast cancer.

## 4. Discussion

Incidence of cancer in Sulaymaniyah city (latitude 33.314690 and longitude 44.376759) in the northeast part of Iraq [21], has lately shown a great increase, as is the case in the rest of Iraq; and breast cancer was the commonest among women [22, 23]. Mitochondrial DNA mutations and polymorphisms have been increasingly reported in a wide variety of cancers, including breast, prostate, and colorectal cancers [24], indicating that mutations and haplogroup determining polymorphisms may influence mitochondrial protein changes that may affect the OXPHOS process and promote the production of reactive oxidative species [25]. For breast cancer it is well established that reactive oxygen species play an important role in the process of carcinogenesis [26, 27]; hence, the current study was conducted to determine common SNPs and haplogroups among the breast cancer cases

**Table 6:** Calculated OR and p values for SNP A8860G with three randomly selected SNPs.

| SNPs | Odd ratio | P value (chi-square) | P value (Fishers exact) |
| --- | --- | --- | --- |
| 8860/750 | 4.722 | 0.0012 | 0.0014 |
| 8860/1438 | 5 | 0.009 | 0.013 |
| 8860/7028 | 5 | 0.011 | 0.021 |

According to the results most of the SNPs were in the coding region, both in the cancer cases and the controls (61% and 58%, respectively). This shows the importance of whole genomic sequencing for precise haplogroup determination and the lesser value of the polymorphisms traditionally used in the hypervariable regions for forensic purposes [28, 29].

Several previous studies have been conducted and showed a significant relation between specific haplogroups and cancer incidence in general [13]. In breast, Chinese women of haplogroups M and subhaplogroup D5 had shown a higher incidence for cancer [30, 31], while no such a remarkable relation was identified between cancer and specific haplogroups in European and Caucasian women [32], still haplogroup K showed a significant association with breast cancer in European-American women [33]. Nevertheless in the current study a significant relation between haplogroup HV and breast cancer was identified with p value = 0.002 and 0.006 for Chi square and Fisher’s exact test respectively and OR of 28.

In addition to the haplogroups, several distinct SNPs have been previously discovered to be associated with cancers in general as T16189C, G10398A and the deletion of mtDNA 4977 [13]; in regard to breast, certain SNPs were identified as well to be associated with increased cancer incidence. A10398G is one of the well-known SNPs in breast cancer detected in European-American, Malaysian and African-American women [33, 34, 35, 36 and 37]; in addition SNPs G9055A and T16519C were also identified as risk factors for breast cancer in European-American females [33]. Furthermore several other germ line mutations as 2463 A-deletion, C6296A, 6298 T-deletion, A8860G, and 8460-13327deletion, were detected in chines women with breast cancer [15].

Although many SNPs were identified in breast cancer samples in the current study shown in (Table 3), but the only mutation showed a significantly high incidence among breast cancer samples compared to the control samples was homoplasmic SNP (A8860G). It is a non-synchronous mutation in the Mt-ATP 6 gene that was detected in all 20 breast cancer samples while only in 4 of the control samples and this result was compatible with Li et al [15]. This gene encodes ATP synthase 6 (681 amino acids), a subunit of complex V, whose mutation results in substitution of a polar uncharged amino acid (threonine) with a non-polar aliphatic amino acid (alanine); this may affect hydrophobic interactions and hence the structure of the protein. However, such a prediction of protein structure is not absolute as these mutations may be followed by other compensatory mutations (suppressor mutations) in order to minimize the initial mutation’s effect [38], these compensatory and suppresser mutations may explain the presence of the mutation A8860G in 20% of phenotypically healthy control samples

## 5. Conclusion

As the results show, both haplogroup HV and SNP A8860G are risky factors for developing breast cancer in the studied population; however these results are not compatible with the previously identified risky SNPs and haplogroups in breast cancer studies performed in other populations, except for SNP (A8860G) which was compatible with that of Li et al [15]; this could be explained by the effect of other parameters on the mitochondrial genome, such as individual physiology and influences of geographical location, suggesting a population specific effect of these haplogroups and SNPs in the carcinogenic processes. Hence in an attempt to fortify the current results and identify possible associations with other tissue cancers, further studies are required with larger sample size and other cancer tissues to be included.

## Supporting information

H and HV calculating table

calculation for mutations (8860) and (750)

Calculation for mutations (8860) and (1438)

calculation for mutations (8860) and (7028)

## Data Availability

The authors confirm that the data supporting the findings of this study are available within the article and its supplementary materials. More details will be provided by corresponding author upon request

## Acknowledgement

We are grateful for the sample donors for their cooperation. Efforts of Mr. Pola Abdalla Othman are gratefully acknowledged. This work was supported by University of Sulaimani and KISSR

## Data availability

All data are available upon request from the corresponding author via the following

## Supporting information

**S1aTable. Frequency difference of haplogroups H and HV among breast cancer samples and control samples**

**S1bTable. Odd ratio for haplogroup HV/H (Risk estimation)**

**S2aTable. Frequency difference of mutations (8860) and (**750**) among breast cancer samples and control samples**

**S2bTable. Odd ratio for 8860/750 (Risk estimation)**

**S3aTable. Frequency difference of mutations (8860) and (1438) among breast cancer samples and control samples**

**S3bTable. Odd ratio for 8860/1438 (Risk estimation)**

**S4aTable. Frequency difference of mutations (8860) and (7028) among breast cancer samples and control samples**

**S4bTable. Odd ratio for 8860/7028 (Risk estimation)**

