## Supplementary material for "Potential Association of Mitochondrial Haplogroups and A8860G Mutation with Breast Cancer Risk": H and HV calculating table

**Supporting information**

**Table S1a**: Frequency difference of haplogroups H and HV among breast cancer samples and control samples

|  | | Group | | Total |
| --- | --- | --- | --- | --- |
|  |  | cancer | control |  |
| Haplogroups | HV | 7 | 3 | 10 |
|  | H | 1 | 12 | 13 |
| Total | | 8 | 15 | 23 |

|  | Value | 95% Confidence Interval | |
| --- | --- | --- | --- |
|  |  | Lower | Upper |
| Odds Ratio for Haplogroups (HV / H) | 28.000 | 2.422 | 323.703 |
| For cohort Group = cancer | 9.100 | 1.326 | 62.462 |
| For cohort Group = Control | .325 | .124 | .849 |
| N of Valid Cases | 23 |  |  |

**Table S1b:** Odd ratio for haplogroup HV/H (Risk estimation)
