## Supplementary material for "Potential Association of Mitochondrial Haplogroups and A8860G Mutation with Breast Cancer Risk": calculation for mutations (8860) and (750)

**Supporting information**

**Table S2a:** Frequency difference of mutations (8860) and (750) among breast cancer samples and control samples

|  | |  | | Total |
| --- | --- | --- | --- | --- |
|  |  | cancer | control |  |
| Site | 8860 | 20 | 4 | 24 |
|  | 750 | 18 | 17 | 35 |
| Total | | 38 | 21 | 59 |

**Table S2b:** Odd ratio for 8860/750 (Risk estimation)

|  | Value | 95% Confidence Interval | |
| --- | --- | --- | --- |
|  |  | Lower | Upper |
| Odds Ratio for Site (8860 / 750) | 4.722 | 1.337 | 16.676 |
| For cohort Group = cancer | 1.620 | 1.121 | 2.342 |
| For cohort Group = control | 0.343 | 0.132 | 0.894 |
| N of Valid Cases | 59 |  |  |
