## Supplementary material for "Potential Association of Mitochondrial Haplogroups and A8860G Mutation with Breast Cancer Risk": Calculation for mutations (8860) and (1438)

**Supporting information**

**Table S3a:** Frequency difference of mutations (8860) and (1438) among breast cancer samples and control samples

|  | |  | | Total |
| --- | --- | --- | --- | --- |
|  |  | cancer | control |  |
| Site | 8860 | 20 | 4 | 24 |
|  | 1438 | 17 | 17 | 34 |
| Total | | 37 | 21 | 58 |

**Table S3b:** Odd ratio for 8860/1438 (Risk estimation)

|  | Value | 95% Confidence Interval | |
| --- | --- | --- | --- |
|  |  | Lower | Upper |
| Odds Ratio for Site (8860 / 1438) | 5.000 | 1.409 | 17.745 |
| For cohort Group = cancer | 1.667 | 1.139 | 2.439 |
| For cohort Group = control | 333 | .128 | .867 |
| N of Valid Cases | 58 |  |  |
