## Supplementary material for "Potential Association of Mitochondrial Haplogroups and A8860G Mutation with Breast Cancer Risk": calculation for mutations (8860) and (7028)

**Supporting information**

**Table S4a:** Frequency difference of mutations (8860) and (7028) among breast cancer samples and control samples

|  | |  | | Total |
| --- | --- | --- | --- | --- |
|  |  | cancer | control |  |
| Site | 8860 | 20 | 4 | 24 |
|  | 7028 | 15 | 15 | 30 |
| Total | | 35 | 19 | 54 |

**Table S4b:** Odd ratio for 8860/7028 (Risk estimation)

|  | Value | 95% Confidence Interval | |
| --- | --- | --- | --- |
|  |  | Lower | Upper |
| Odds Ratio for Site (8860 / 7028) | 5.000 | 1.376 | 18.168 |
| For cohort Group = cancer | 1.667 | 1.117 | 2.487 |
| For cohort Group = control | .333 | .127 | .874 |
| N of Valid Cases | 54 |  |  |
